# Investigating the Y chromosome in complex disease: Phenome-wide scan across 104,334 Finnish men

**DOI:** 10.64898/2026.06.09.26355235

**Authors:** Annina Preussner, Jaakko Leinonen, FinnGen, Aoxing Liu, Matti Pirinen, Taru Tukiainen

**Affiliations:** Institute for Molecular Medicine Finland (FIMM), Helsinki, Finland; Center for Child, Adolescent, and Maternal Health Research, Faculty of Medicine and Health Technology, Tampere University, Tampere, Finland; Analytic and Translational Genetics Unit, Massachusetts General Hospital, Boston, MA, USA; Program in Medical and Population Genetics, Broad Institute of MIT and Harvard, Cambridge, MA, USA; Center for Genomic Medicine, Massachusetts General Hospital, Boston, MA, USA; Stanley Center for Psychiatric Research, Broad Institute of MIT and Harvard, Cambridge, MA, USA; Department of Public Health, Faculty of Medicine, University of Helsinki, Helsinki, Finland; Department of Mathematics and Statistics, University of Helsinki, Helsinki, Finland; Institute of Clinical Medicine, School of Medicine, Faculty of Health Sciences, University of Eastern Finland, Kuopio, Finland

## Abstract

Although the Y chromosome represents roughly 2% of the male genome, it is often ignored in genome-wide association studies (GWAS). Subsequently, the potential health impacts of Y-chromosomal genetic variation remain incompletely understood. To fill this gap, we performed a phenome-wide association study (PheWAS) in FinnGen across 1,426 binary and quantitative traits using Y-chromosomal variation (frequency ≥ 1%) in 104,334 genotyped men. As Y chromosome variation is prone to population stratification, we performed carefully adjusted association analyses and further examined these through kin-based validation in 19,275 female and 24,712 male 1^st^ degree relatives. We found 121 suggestive (p < 5.6x10^-3^) phenotypic associations in the Y chromosome, yet none of these were strong enough to reach phenome-wide significance (p < 3.9x10^-6^). While only 38 associations were supported in the kin-based validation, intriguingly we found support for a previously suggested link between haplogroup I1 and coronary heart disease (CHD; OR=1.06, 95%CI=1.02-1.11, p=3.7x10^-3^; male validation OR=1.05; female validation OR=0.97). The I1-CHD association was detected across distinct geographical areas within Finland and was independent from Loss of Y (LOY) and the autosomal risk to CHD, proposing a link between germline Y-chromosomal variation and heart disease risk. Overall, this study presents a comprehensive phenome-wide analysis of Y-chromosomal associations, highlighting the potential relevance of Y-chromosomal variation beyond sex determination. Our findings further emphasize the need for improved capture of Y-chromosomal variants and further analyses in biobank-scale data to allow for deeper exploration of male-specific genetic architecture of complex diseases.

**GRAPHICAL ABSTRACT:** 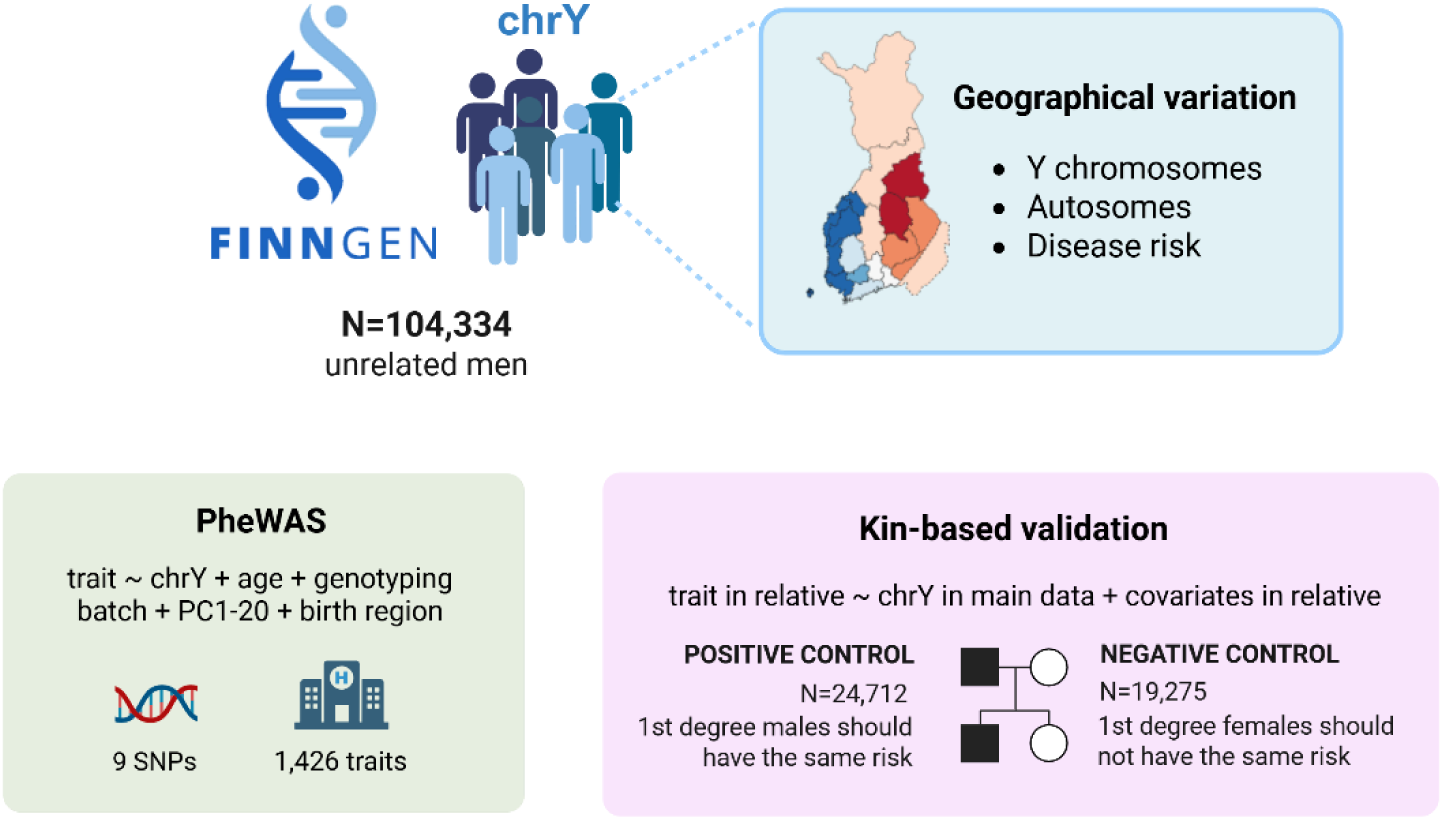

**Graphical Abstract** Overview of the study. Using a dataset of 104,334 unrelated genotyped Finnish men we performed a PheWAS across 1,426 binary and quantitative traits and common Y-chromosomal variation (MAF ≥ 1%), adjusting for geographical confounding factors known to vary within the population. To confirm suggestive associations identified in the PheWAS, we performed kin-based validation to infer whether the association is likely linked to the Y chromosome, using the samples’ 1st degree male (positive control) and 1st degree female (negative control) relatives’ phenotype data.

## INTRODUCTION

Over the past decades, genome-wide association studies (GWAS) have greatly advanced our understanding of the genetic basis of complex traits^1^. However, the Y chromosome is commonly excluded from such studies, making it one of the most understudied regions in complex trait genetics^2–4^. This is largely due to its unique characteristics, which pose challenges for standard GWAS approaches. In particular, the lack of recombination across most of the Y chromosome results in strong correlations between the variants and pronounced population stratification, complicating the identification of independent association signals. Additionally, its haploid nature requires modelling strategies to properly account for allele dosage, and the inclusion of only males in such analyses further limits the power to detect statistically meaningful associations.

Somatic loss of the Y chromosome (LOY) in blood cells has provided compelling evidence that Y chromosome influences male health well beyond sex determination and fertility. LOY has been associated e.g., with increased risk of cardiovascular disease, type 2 diabetes (T2D), cancer, and reduced lifespan^5^, all characteristics that show sex differences, proposing that perturbation of Y-chromosomal content can have systemic consequences. In contrast, the contribution of inherited (germline) Y-chromosomal variation to complex traits remains poorly understood. Despite the presence of broadly expressed genes and regulatory elements on the Y chromosome^6,7^, germline variation has rarely been examined in large-scale association studies, leaving its potential role in male-biased disease risk largely unexplored.

While Y chromosome has been included in individual association studies for reproductive disorders^8^, cancer^9,10^, behavioral traits^11^, infectious diseases^12^, neurodevelopmental and neuropsychiatric disorders^13,14^, cardiometabolic traits^15–18^, the findings have remained inconsistent and difficult to interpret. While some associations have been reported, limited statistical power or potential confounding by population stratification remain as common issues for many Y chromosome studies. For instance, Eales et al. (2019) reported a positive association between Y-chromosomal haplogroup I1 and atherosclerosis in the UK population, yet later Timmers and Wilson (2022) suggested this association to be driven largely by population stratification. Moreover, as most of the well-powered studies have focused merely on Y chromosomes from the UK and Japan^16–18^, much of the Y chromosome’s genetic diversity has left unexplored. Consequently, diverse populations and a broader range of phenotypes are needed to obtain a better understanding of the Y chromosome’s potential contribution to complex disease.

In this study, we performed a PheWAS between 1,426 traits and Y-chromosomal variants among 104,334 men from FinnGen, providing an ideal framework for investigating both regionally restricted haplogroups (N1a1) and more widespread ones (I1, R1a, R1b) within a single population. We assessed the robustness of the associations by kin-based validations and geographically stratified analyses. Taken together, our findings as well as careful methodological considerations provide a foundation for further analyses of Y chromosome variation at the biobank-level and highlight the need for enhanced capture of Y-chromosomal variants to facilitate these efforts.

## MATERIALS AND METHODS

### Genotyping array data QC

For this study, we utilized the FinnGen project^19^ release 12 data, comprising 500,348 samples with detailed health registry data. The details on genotyping have been described previously^19^. To ensure consistency and reduce bias (**Supplementary Methods**), we analysed 140,234 males genotyped on the FinnGen custom array v2, including 554 Y-chromosomal markers before quality control (aligned to GRCh38).

For quality control, we set all heterozygous calls missing and removed SNPs with more than 10% missingness. We then excluded samples with more than 10% missingness or classified as having non-Finnish genetic ancestry based on PCA and kept only samples that had covariate data available (age, autosomal genotype PCs 1-20, birthplace and genotyping batch). Additionally, we removed samples with Klinefelter syndrome (47, XXY) based on a clinical diagnosis, and samples for which we could not determine a haplogroup with Yhaplo^20^ due to contradicting SNPs. We also removed closely related samples (KING^21^ kinship coefficient > 0.125), yielding a sample of N=104,334 unrelated men.

Next, we kept variants with at least 1% frequency in the data, yielding in 60 Y-chromosomal SNPs. We used the YFull^22^ and ISOGG^23^ to annotate the haplogroup-defining status of the SNPs (**Suppementary Methods**). Of the identified nine haplogroup clades from our data (I1, N1a1, R1a, R1b, IJ, K, P1, I1a2-Z73, R1a-Z284) (**Table S3**), we selected a single SNP per haplogroup for the association analyses as the variants within each haplogroup were fully correlated. Details of each step are listed in **Table S1**.

### Exome sequencing data QC and annotation

To assess if rare variants were enriched to certain haplogroups, we investigated whole exome-sequencing (WES) data, which had been performed for a subset samples part of FinnGen. After quality control and annotation (**Supplementary Methods**) we had 9,736 samples and 300 variants in the data.

### Phenotype selection

For the association testing, we selected clinical diagnoses and phenotypes (i.e., endpoints) defined by FinnGen having at least 100 cases among the samples. After removing highly correlated phenotypes^24^ we ended up with N=1,413 binary endpoints. We further selected 13 quantitative endpoints including height, weight, number of offspring, and 10 common laboratory measurements into the analyses (**Supplementary Methods**; **Table S2**).

### Loss of Y chromosome (LOY)

Further, we assessed whether LOY occurrence differed between haplogroup carriers. The LOY status was available for FinnGen R9 data, previously defined using MoChA^25,26^, with GRCh38 as the reference (see e.g., Koskela et al. 2021; Zekavat et al. 2021). Using these LOY calls, we fitted a logistic regression model with LOY status as the outcome and haplogroup (a categorical variable with haplogroups N1a1, I1, R1a, R1b, I-Z73, R-Z284, and ‘other’ corresponding to all remaining haplogroups) as the predictor, adjusting for age at sample collection, genotyping batch, PCs 1-20 and birth region as covariates in R (version 4.5.0)^29^. Overall haplogroup effects were evaluated using a likelihood ratio χ^2^-test based on the logistic regression model deviance (df = 6). Age-adjusted LOY prevalence estimates for each haplogroup were obtained using estimated marginal means^30^. We compared the rate of LOY occurring in at least 10% of the cells (defined for N=56,715 samples), as well as without a specified threshold (defined for N=64,150 samples).

### Association testing

The association analyses were performed with REGENIE^31^, since it is commonly used for association analyses of the whole genome. As the analyses were focused on the haploid Y chromosome, we omitted step 1 in REGENIE, which adjusts for autosomal structure and relatedness, because it does not account for Y chromosome inheritance. Rather, to control for relatedness, we excluded closely related samples. For the regression models (i.e., step 2) we chose age, genotyping batch, PCs 1-20 and birth region as covariates.

Because REGENIE applies diploid (0/2) rather than haploid (0/1) genotype coding for the Y chromosome, the estimated effect sizes and standard errors are half of their haploid values (**Figure S2**). To report the effects on the haploid scale, all β coefficients and corresponding SEs from REGENIE were multiplied by two. The 95% CI was calculated as

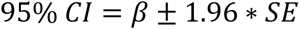

where β for logistic regression was on the log-odds ratio scale.

### Significance threshold and power calculations

The PheWAS significance threshold was determined using Bonferroni correction (p=0.05/9/1426=3.89x10^-6^), and using this threshold, we performed statistical power analyses for each haplogroup (**Supplementary Methods**). We also applied a more liberal p-value threshold (p=0.05/9= 5.5x10^-3^) to flag associations as suggestive.

### Kin-based validation - Effect size comparison between first degree relatives

To validate the suggestive associations, we compared effect sizes of the Y chromosome variant carriers to those of their 1^st^ degree relatives under a kin-cohort design^17,32^. Because first-degree male relatives share the same Y chromosome, their effect sizes should be similar for a given Y-linked association, whereas in female relatives the effect should attenuate. For estimating the male relatives’ effects (positive control) we used N=24,712 samples and for female relatives’ effects (negative control) we used N=19,275 samples (**Supplementary Methods**; **Table S2**). The association analyses were performed in REGENIE^31^ by using the Y-chromosomal genotype of the sample itself, while replacing the phenotype data (disease status and covariates) from the relative. All suggestive associations with at least five carriers in the kin-based validation sub-datasets were analysed.

To evaluate concordance between the main analysis effect estimate and the kin-based validation effect, we applied linemodels^33^ in R (version 4.5.0)^29^. To probabilistically estimate whether the same effect was present in both analyses, we applied two linemodels with equal priors: “M1” to model similar effects, where β_main_ = β_kin_ (slope=1), and “M0” to model no effect in the kin analysis, where β_kin_= 0 (slope=0). We classified associations as concordant or discordant based on whether the kin-validation models showed support for association in the same direction (M1) or supported the null model (M0), using a posterior probability threshold of PP > 0.5 (**Supplementary Methods**).

### Assessing effects between haplogroup pairs

Because the effect size of a given SNP or haplogroup depends on the reference group used for comparison, which varies across populations, we conducted additional analyses evaluating each haplogroup’s effect relative to other haplogroups (**Supplementary Methods**).

### Adjusting for possible CHD confounders

To determine whether the association between Y-chromosomal variation and CHD could be explained by confounding factors, we performed several additional association analyses implemented in R (version 4.5.0)^29^. We performed separate logistic regression analyses adjusting for risk factors, with current smoking status (yes/no; N=64,244) and LOY status (yes/no; N=64,249), considered in separate models. To assess whether the CHD association is attributable to autosomal rather than Y-chromosomal variation, we incorporated a polygenic risk score (PRS) for CHD into the analysis (**Supplementary Methods**). First, we added the PRS as a covariate into the logistic regression and then further performed stratified analysis for high (top 10%), intermediate (10-90%) and low (bottom 10%) PRS groups. To assess whether Y-chromosomal variation contributes independently to CHD risk, we performed an association test for CHD excluding the Y chromosome variant (while retaining all other covariates) and compared the models using a likelihood-ratio test. To account for potential geographical confounding, we conducted stratified analyses across distinct regions (**Supplementary Methods**).

## RESULTS

### FinnGen genotype data identifies major Y-chromosomal haplogroups but provides limited sublineage resolution

We analyzed common variants (≥1% frequency) in the male-specific region of the Y chromosome (MSY) among 104,334 unrelated men in FinnGen (**Figure 1A**). From the initial 554 Y-chromosomal markers on the genotyping array, 10% (N=58) passed quality control criteria and were polymorphic among the participants (**Table S1**). Owing to the MSY presenting a single haplotype and many variants being correlated, we focused on a set of nine variants corresponding to distinct lineages on the Y-chromosome phylogeny (**Figure 1A**). These variants captured the four major haplogroups in Finland, N1a1 (61% of the samples), I1 (26%), R1a (6%), and R1b (5%), with similar frequencies as previously described^34^. In addition, we identified markers for three upstream haplogroups P1 (11%), K (72%) and IJ (27%), and two downstream haplogroups I1a2-Z73 (1%) and R1a1-Z284 (2%) (**Figure 1A; Figure S1**).

We next sought to quantify how well the standard European genotyping array used captures Finnish Y-chromosomal variation. Utilizing the number of common downstream lineages from our previous study^34^, we quantified that 98/100 sublineages in Finland (i.e., partially overlapping clades within the Y-chromosome phylogeny) were not captured with the genotyping array (**Figure 1A**). For instance, while haplogroup N1a1 divides into lineages N-VL29 and N-Z1934, that display highly distinct geographical enrichment patterns^34^ neither of these lineages, nor any of their sublineages were captured in the data (**Figure 1B; Table S3**). Similarly, for I1, R1a, and R1b, we failed to capture more detailed variation, apart from the two downstream lineages I1a2-Z73 and R1a1-Z284 (**Figure 1A**).

**Figure 1.**
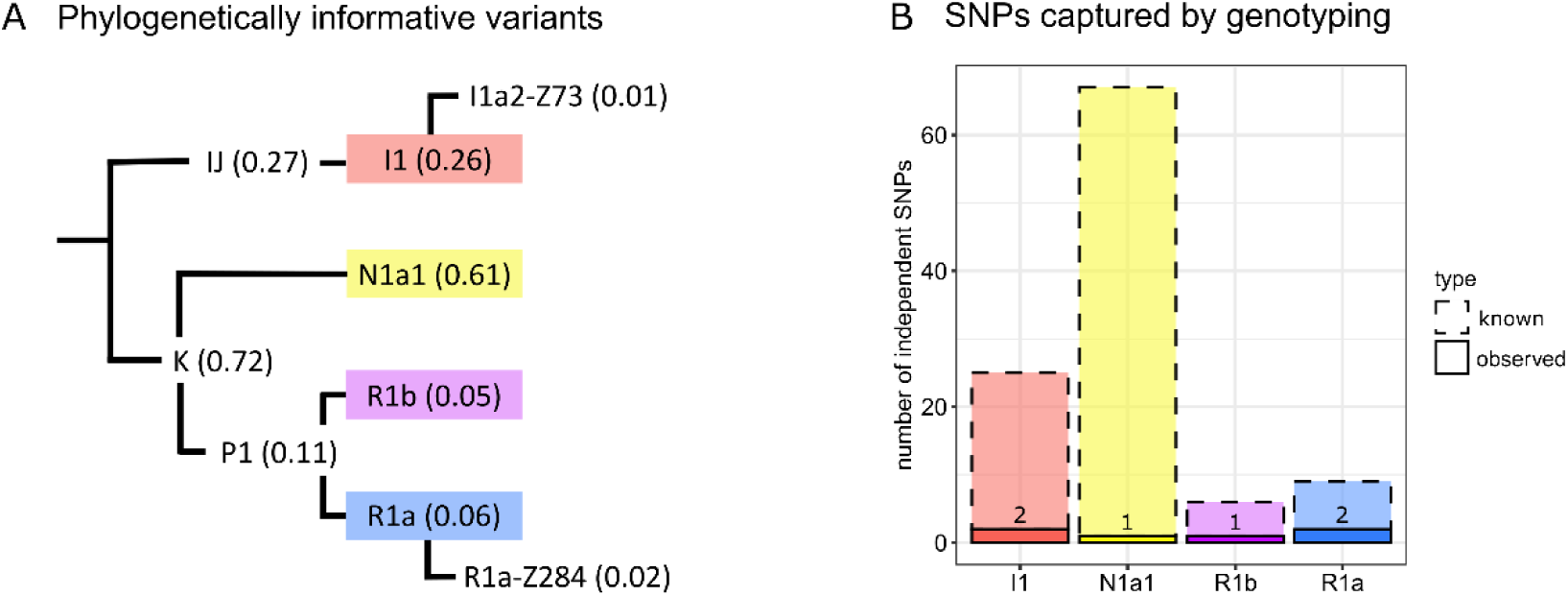
Y-chromosomal haplogroups captured in FinnGen. A) Phylogenetic presentation of the studied Y-chromosomal haplogroups and their relationships, with the observed frequencies in parentheses. The four major Finnish haplogroups N1a1, I1, R1b and R1a are highlighted with colors. B) Comparison of the number of observed SNPs defining sublineages within each major haplogroup versus the number of SNPs defining known sublineages (i.e., partially overlapping clades within the Y-chromosome phylogeny) for each haplogroup, present at a frequency of at least 1% frequency in the Finnish population^34^.

### Y-chromosomal variation in FinnGen shows suggestive associations for complex diseases

To elucidate the potential impact of Y-chromosomal variation on complex disease, we performed a PheWAS between the nine Y-chromosomal variants and 1,426 traits among 104,334 unrelated men (**Figure 2; Table S4**). For common binary traits such as asthma (with ∼10,000 cases), our data provided sufficient power (80%) at the PheWAS threshold (p=3.9x10^-6^) to detect odds ratios (OR) starting from 1.13, depending on the frequency of the variant (**Figure S3**).

We applied two significance thresholds in the PheWAS. Using a conservative multiple-testing correction p<3.9×10⁻⁶, no significant associations were observed (**Figure 2; Table S4**). With the more liberal threshold (p<5.6×10⁻³), we observed in total 121 associations, termed as suggestive, spanning through various disease categories (**Table S4**). As no associations were observed for traits with a strong socioeconomical or regional component (e.g., number of children or medication intake), we consider the association models properly adjusted for geographical confounding.

For haplogroup I1, previously implicated in the UK population for cardiometabolic traits such as coronary heart disease^16,17^ and type 2 diabetes^18^, we observed 18 suggestive associations, with many of these enriched for cardiovascular traits (e.g., hard cardiovascular diseases OR=1.06, 95%CI=1.02-1.10, p=2.9x10^-3^) and traits related to cardiovascular disease, such as type 2 diabetes (OR=1.06, 95%CI=1.02-1.10, p=2.2x10^-3^) and haemoglobin A1c levels (β=0.03, 95%CI=-0.11-0.08, p=1.6x10^-3^) (**Figure 2; Table S4**). For haplogroup I1a2-Z73 we observed 16 suggestive associations, mostly related to musculoskeletal disorders and neoplasms. Haplogroup R1a1-Z284 displayed 10 suggestive associations across various traits.

Notably, the fewest suggestive associations were observed for haplogroups N1a1 and R1a (**Figure 2**). For haplogroup N1a1, we observed nine suggestive associations of which the majority were mainly risk-decreasing (**Figure 2; Table S4**). For haplogroup R1a we observed four suggestive risk-increasing associations related to eye, infectious and digestive traits (**Figure 2; Table S4**). Finally, for haplogroup R1b we observed 18 suggestive associations, with several related to colorectal neoplasms (**Table S4**).

As robustness analysis we analyzed the effects of the upstream lineages P1, K and IJ (**Figure 1**; **Table S5**). For instance, since IJ and I1 capture largely the same set of individuals within the Finnish population (**Figure 1**), reassuringly all the suggestive associations observed for IJ reflected the associations of I1 (**Figure 2**; **Table S5**). Moreover, because haplogroups K (upstream of N1a1 and R) and I1 are mutually exclusive (**Figure 1A**), the associations observed for I1 were mirrored in the opposite direction for haplogroup K. On the other hand, haplogroup P1, which combines carriers of R1a and R1b, showed mostly attenuated effects compared to R1a or R1b (**Table S5**).

**Figure 2.**
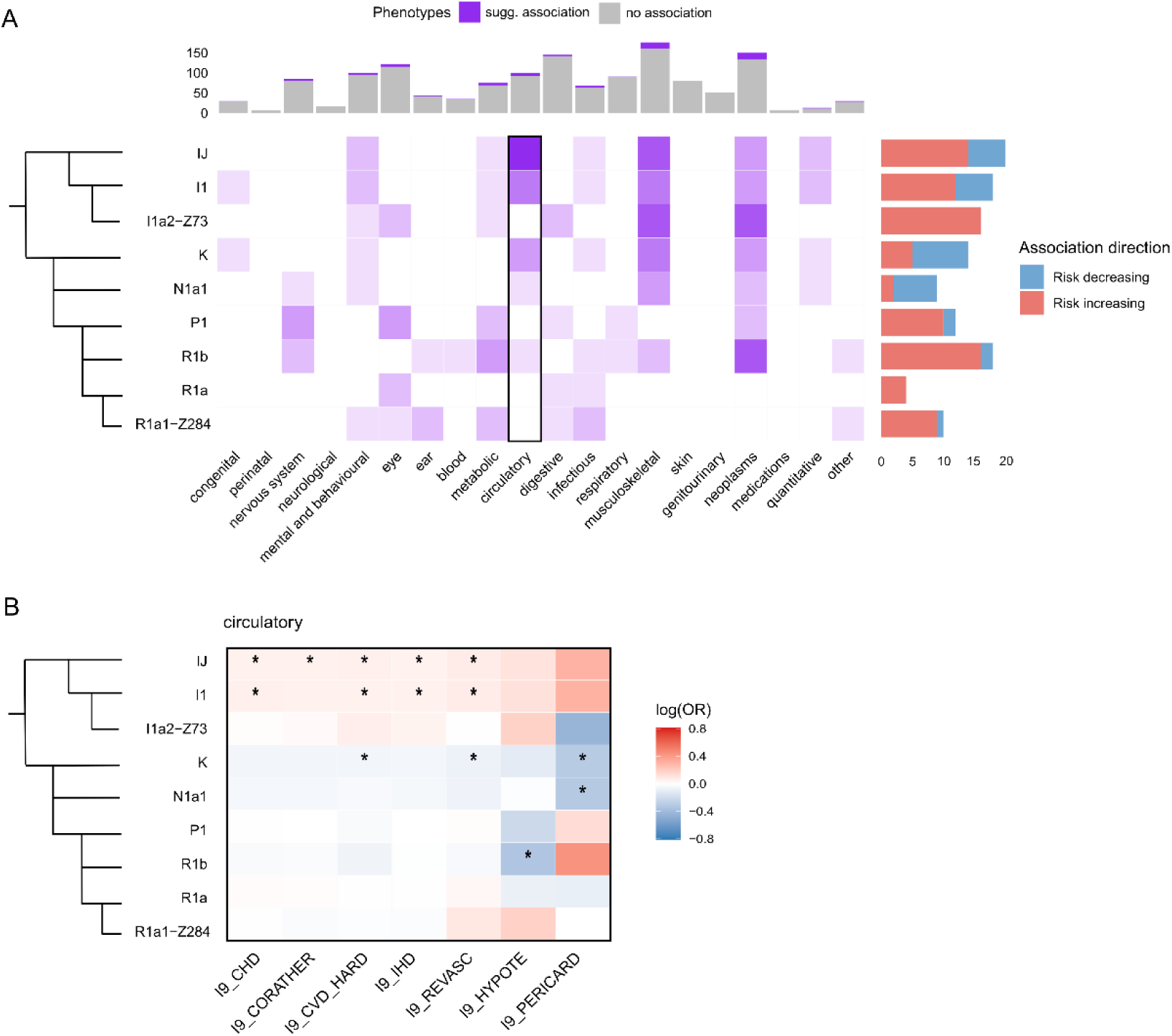
PheWAS results for A) all suggestive associations (p < 0.05/9) of Y-chromosomal haplogroups in FinnGen. The color intensity on the heatmap indicates the number of suggestive associations per haplogroup and phenotype category. The barplot on the right indicates the number of risk-increasing (red) and risk-decreasing (blue) associations per haplogroup. The barplot on the top indicates number of tested phenotypes in each category and amount of suggestive associations in the category. The simplified dendrogram on the left indicates the approximate relationships of the haplogroups. B) A heatmap of circulatory phenotypes that show suggestive associations. Suggestive associations are marked with a star and the color indicates the strength of the association on log(OR) scale.

### Kin-based validation supports several suggestive associations

To follow-up the suggestive associations identified in the PheWAS, we utilized the extensive set of related samples in FinnGen to conduct kin-based validation. Briefly, we compared the PheWAS effect estimates to associations between each haplogroup and the phenotypes of first-degree male and female relatives (**see Methods**) (**Figure 3A-B**; **Table S5**). Overall, 32% of the associations followed-up in the kin-based validation displayed concordant effects (38/120), meaning the effect from the PheWAS analysis was supported by observations in the male validation, while the female validation supported a null effect, as expected for a true Y-chromosomally driven association (**Table S5**). Among the major haplogroups, I1 and R1b displayed the highest number of concordant effects in the validation (38% and 44%, respectively). The concordant effects for haplogroup I1 were predominantly associated with cardiovascular traits (**Figure 3**; **Table S5**) whereas for R1b they were mainly linked to colorectal neoplasms (**Table S5**). For instance, the effect between I1 and CHD (OR=1.06, 95%CI=1.02-1.10) was more similar to the effect observed in the male validation (OR=1.05, 95%CI=0.95-1.15, PP=0.57), than in the female validation (OR=0.97, 95%CI=0.84-1.13, PP=0.36) (**Figure 3C**; **Table S5**).

For the remaining 82 suggestive associations, the kin-based validation did not support Y-chromosome driven effects. Here, the PheWAS effect was either concordant with the effect from female validation only (N=28), concordant with both male and female validations (N=10), or neither (N=44) (**Table S5**). This suggests the original PheWAS result may partly reflect shared socioeconomic conditions, lifestyle and environmental factors or statistical fluctuation. For instance, the associations between T2D and I1 was not concordant with the original result (OR=1.06, 95%CI=1.02-1.10) either in male (OR=1.02, 95%CI=0.94-1.10; PP=0.39) or female (OR=1.03, 95%CI=0.93-1.14, PP=0.48) relatives (**Figure 3E**).

**Figure 3.**
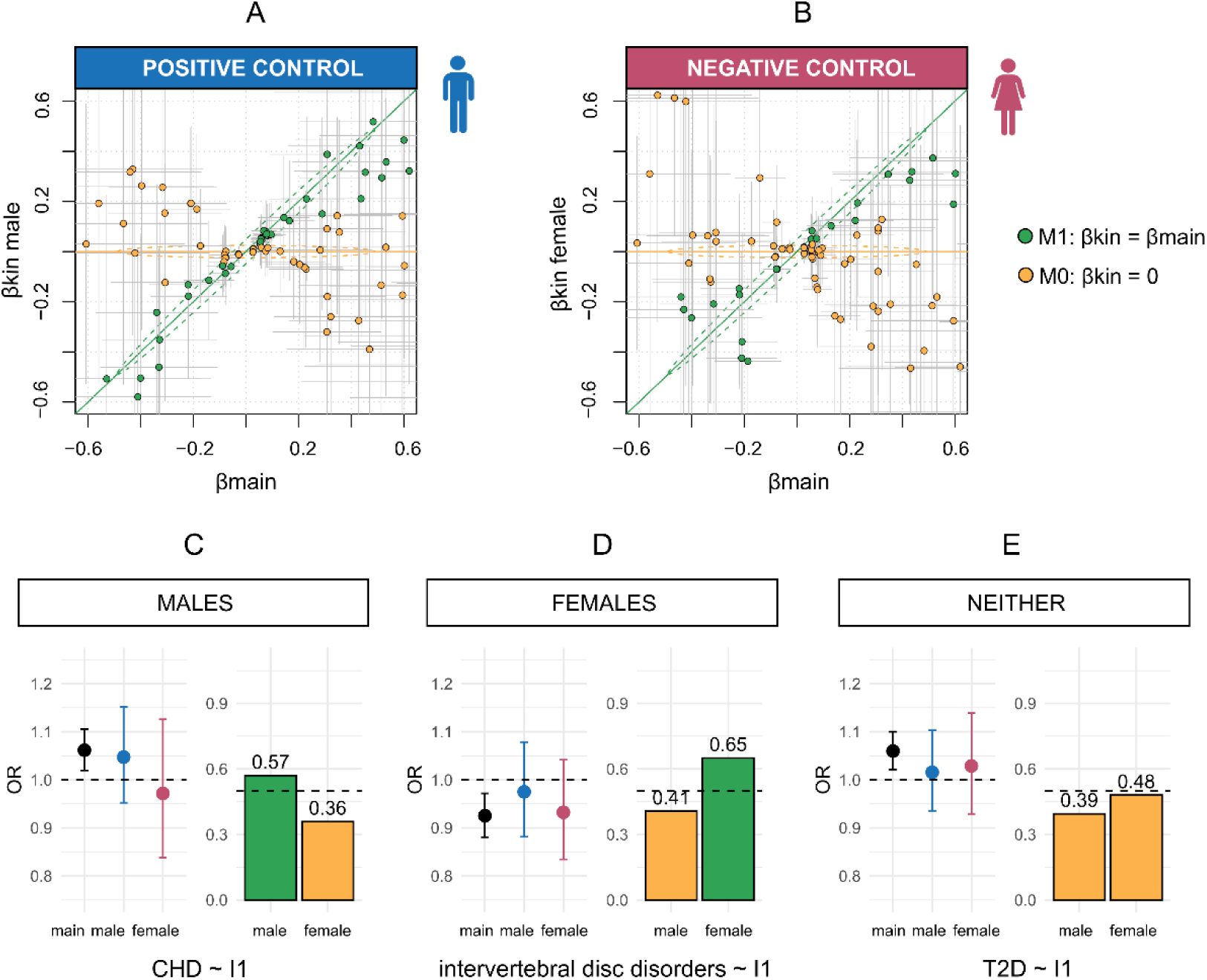
Kin-based validation for suggestive Y-chromosomal associations in FinnGen. The main (PheWAS) effect was compared to A) male (N=24,712) and B) female (N=19,275) relative’s effects with linemodels^33^. The green color corresponds to the effect being concordant between the main analysis and the kin-based validation with posterior probability (PP) > 0.5 for model M1 (βkin = βmain), while the orange color corresponds to the effect being discordant between the main analysis and the kin validation with PP > 0.5 for model M0 (βkin = 0). C) An example of a suggestive association with a concordant effect in males, visualizing the OR and 95% CI observed for CHD in all three association analyses and the PPs for the model M1 in both kin-based validations. Black color corresponds to main PheWAS effect, blue to the male validation effect and pink to the female validation effect. On the barplot, the bars crossing the M1 PP>0.5 are colored with green, corresponding to concordant effects, while bars below the threshold are colored with orange, corresponding to discordant effects. Panels D-E visualize examples of effects that D) are concordant in females for intervertebral disc disorders and E) an association that is not concordant in either of the relatives for T2D.

### Association between cardiovascular disease and haplogroup I1

Previous studies in the UK population have suggested a link between haplogroup I1 and CHD^15–17^, with effect estimates ranging from a notable risk (OR=1.11, 95%CI=1.04-1.18, p=6.8x10^-4^)^16^ to a marginal association (OR=1.06, 95%CI=1.00-1.12, p=0.058)^17^, leaving uncertainty as to whether this reflects a true genetic effect or residual population stratification. In Finland, this concern is particularly relevant given pronounced geographical stratification of both Y-chromosomal haplogroups and CHD: haplogroup I1 is enriched in southwestern Finland (SW) (**Figure 4A**), whereas CHD prevalence and polygenic risk are higher in the northeast (NE)^35,36^ (**Figure S4A, Figure 4B**). We therefore examined the extent to which the I1-CHD association in our study could be explained by confounding factors.

In regionally stratified analyses, we did not observe significant differences in I1 effects between NE (OR=1.08, 95%CI=1.01-1.15, p=0.031; N=40,328; I1 frequency 19%) and SW (OR=1.05, 95%CI=1.01-1.10, p=0.051; N=62,834; I1 frequency 31%) (z-test for effect size difference p=0.48) (**Figure 4C**; **Table S6**). Similarly, no significant differences in effect sizes were observed between the more detailed 20 sub regions (Q-test for heterogeneity p=0.75) (**Figure S4B**).

The I1 effect remained consistent also after including the CHD PRS in the association model (OR=1.06, 95%CI=1.02-1.10, p=7.1x10^-3^) (**Figure 4B;4D**), and when limiting the analysis to high-risk individuals (top 10% of the CHD PRS) (OR=1.09, 95%CI=0.97-1.23, p=0.14) (z-test for effect size difference compared to the full sample p=0.61) (**Table S6**). Furthermore, adding haplogroup information in the model significantly improved model fit for predicting CHD compared with a model containing only age, birth region, PCs, and batch (Likelihood-ratio test p=4.0x10^-3^), suggesting that I1 may act as an independent predictor of CHD risk from the available covariates.

Adjusting the association model with additional risk factors, such as smoking or LOY, did not impact the observed effect (smoking adjusted OR=1.06; 95% CI=1.00-1.12, p=0.053; LOY adjusted OR=1.05; 95% CI=1.00-1.10, p=0.071) (**Figure 4D**). Finally, we assessed whether changing the reference haplogroup affected the results and found that it had no significant impact, with I1 showing an increased risk for CHD in all comparisons (OR=1.02-1.07) (**Table S7**).

**Figure 4.**
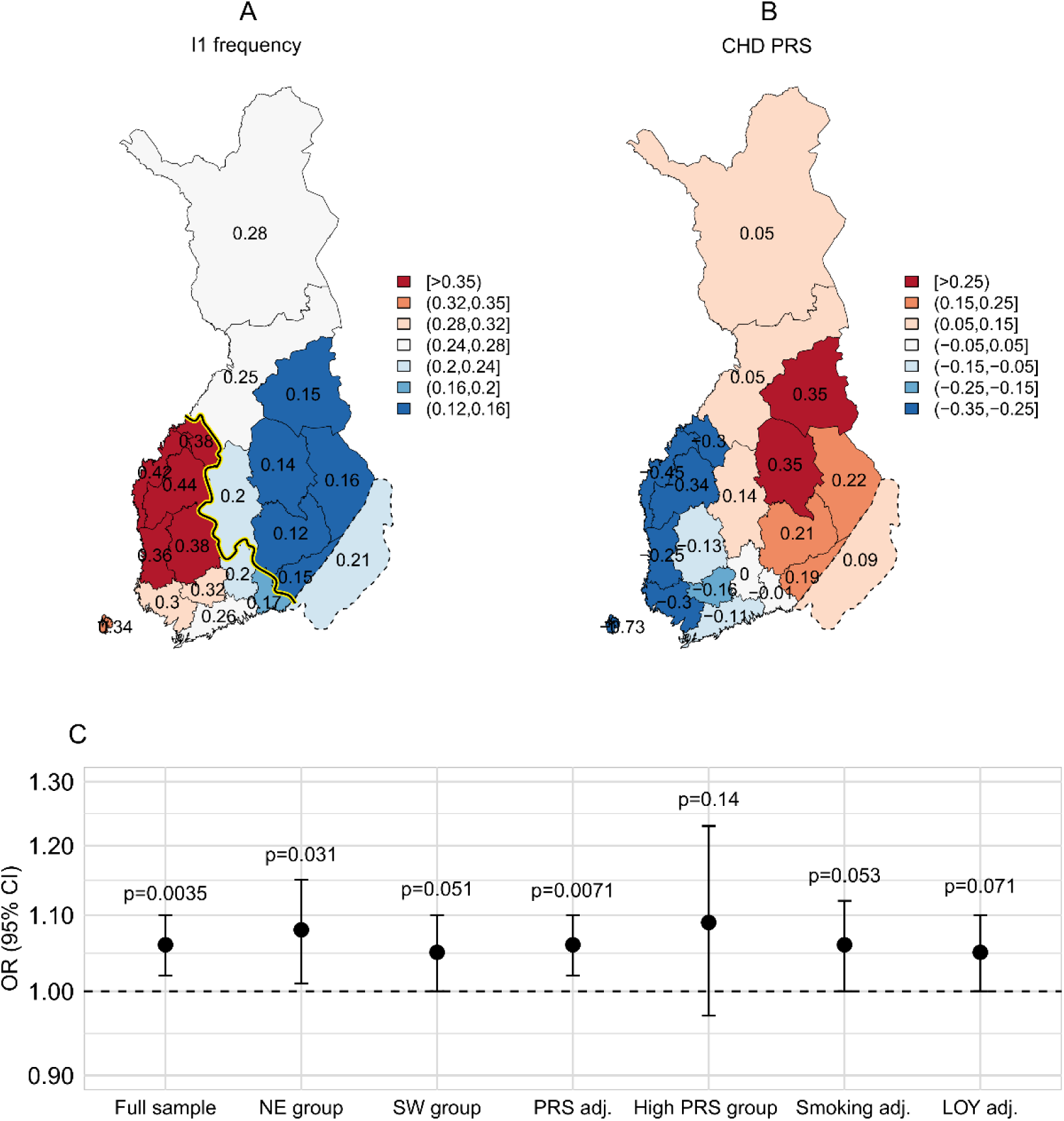
Detailed evaluation of confounding in the relationship between I1 and CHD. A) Geographical distribution of haplogroup I1, and B) the geographical distribution of CHD PRS among 20 Finnish regions. Panel A uses red and blue shading to indicate I1 frequencies above and below the overall average of I1 frequency across the country, respectively, while in panel B red and blue shading represent higher and lower PRS values compared to the average across the country, respectively. The division between Northeast (NE) and Southwest (SW) regions is highlighted in panel A with a yellow line. C) Results for the CHD-I1 association in the full sample compared to association results from further models, by stratifying the samples by region (NE group, SW group), the top 10% of autosomal CHD PRS (high PRS group), and by adjusting the full sample for possible confounders (autosomal PRS, Smoking status, and LOY status).

### LOY and protein-coding variation on Y chromosomes

Identifying causal variation for Y chromosome associations is difficult as the variants reside on a single haplotype and thus are in complete linkage disequilibrium. To explore factors that might underlie the associations, we examined the enrichment of both somatic (i.e., LOY) and rare coding Y-chromosomal variation in each haplogroup. Although strongly deleterious variants would be unlikely enriched to a Y-chromosomal haplogroup, variants with mild, context-dependent, or late-onset effects that do not substantially reduce male reproductive fitness may persist and drift to substantial frequencies on the Y chromosome^37^.

The age-adjusted prevalence of LOY was on average 15.9% and 6.9% for any LOY and large LOY (>10% of the cell fraction exhibiting LOY), respectively, across the carriers of haplogroups N1a1, I1, R1a, R1b, I1-Z73, R1a-Z284, and other (**Table S8**). There were no significant differences between LOY prevalence between the haplogroups (χ^2^ test any LOY p=0.05; large LOY p=0.06) (**Table S8**; **Figure S5**).

In the WES data, we identified 300 variants across 45 genes on the Y chromosome, of which 31 were annotated as missense and seven as loss-of-function variants (protein-altering variants, PAV). After normalizing the number of PAV observed in each major haplogroup, we observed that the number of these variants was highly variable between haplogroups (χ^2^ test for equality of proportions, p=1.43x10^-8^). Most notably, haplogroup N1a1 had fewer PAV per sample (average 0.004) compared to other haplogroups (ranging from 0.019 to 0.022 per haplogroup) (**Table S9**).

Although haplogroup I1 did not show an enrichment for any given type of variant, we observed a gene-level enrichment for missense variants in *KDM5D* in I1 carriers. Intriguingly, among 20 samples with a detected mutation in this gene, 18 were attributed to a single variant chrY:19706254 A>G, which is annotated as a Finnish-enriched variant in gnomAD^38^. Although none of the carriers of this variant had a CHD diagnosis, the relatively young age of the I1 carriers in this data (mean age 44) makes it possible that some may develop CHD later in life. Consequently, based on this data we cannot conclude whether this variant explains the relationship between I1 and CHD observed in FinnGen.

## DISCUSSION

Here, we conducted a PheWAS for Y chromosome variation including 1,426 traits and 104,334 genotyped Finnish men. Importantly, we were able to include the haplogroup N1a1 for the first time into a well-powered association analysis, along with haplogroups IJ, I1, K, P, R1a, R1b, I1a2-Z73 and R1a1-Z284. We found suggestive evidence for the role of germline Y-chromosomal variation in diverse complex disease phenotypes, which we evaluated further by rigorous downstream analyses. Notably, we observed support for the previously speculated link between haplogroup I1 and cardiovascular disease (CHD; OR=1.06, p=3.5x10^-3^). To continue exploring these associations, future efforts should focus on capturing a broader range of common and rare Y-chromosomal variants. Our study further highlights that interpreting Y-chromosomal results require careful consideration of methodology, statistical power, and multiple-testing constraints.

Despite the absence of phenome-wide or genome-wide significant associations in our study, we identified 121 suggestive associations (p < 0.05/9). To assess the robustness of these suggestive associations, we leveraged the extensive relatedness in FinnGen for a kin-based validation. In the PheWAS the effects sizes for I1 in relation to CHD and T2D were comparable to those previously reported in the UK population^16–18^. However, kin-based validation supported only the association with CHD. Notably, the I1-CHD effect estimate was more consistent with the conservative UK estimate, derived from models rigorously adjusted for population stratification^17^. Replication of the I1-CHD association in Finland is particularly informative given the distinct haplogroup composition relative to the UK population^16^, suggesting that the observed associations may reflect a shared underlying genetic source. Further analyses indicated that the association was not explained by geography, autosomal CHD risk, or conventional factors such as smoking. Adjustment for LOY, which has been linked to heart disease^39^, had no effect, supporting the interpretation that somatic and germline Y-chromosomal variation may independently contribute to CHD risk.

Beyond haplogroup I1, we identified several suggestive associations for other haplogroups on phenotypes many of which have not been previously studied in the context of the Y chromosome. For instance, haplogroup R1b was linked with colorectal conditions that are known to be more prevalent in males^40^, such as adenocarcinoma of rectum (OR=1.62, p=6.6x10^-4^), which also showed concordant effects in the kin-based validation (males OR=1.52; females OR=0.63). These associations, provided validation in other data sets, can help elucidate male-specific disease risks mechanisms.

Notably, we observed few suggestive associations for haplogroup N1a1, regardless of its high frequency in the Finnish population and most of the suggestive associations for N1a1 were risk-decreasing. A depletion of N1a1 associations may partly reflect the observation of N1a1 carriers showing overall fewer Y-linked LoF and missense variants compared to other haplogroups. Additionally, importantly, the absence of associations could stem from our inability to resolve N1a1 into more specific haplogroups, including the two distinct N1a1 lineages common in Finland^34^.

Overall, our PheWAS was limited to detecting effects at main Y-chromosomal haplogroup level. The genotyping array used in FinnGen captured only nine phylogenetically distinct variants, as the bulk of the Y-chromosomal markers on the array were rare or not detected in Finns, these rather reflecting variation in men of non-Finnish European ancestry. Overall, this highlights that genotyping array performance for Y-chromosomal analyses is substantially reduced when the population context is not incorporated into array design, underscoring the need to optimize variant capture for the target population.

The unique inheritance pattern of the Y chromosome poses specific challenges for association analyses and interpretation that are not encountered in standard GWAS frameworks. Nevertheless, our results demonstrate that Y-chromosomal variation can be analysed using conventional GWAS tools, provided that chromosome-specific assumptions are carefully considered and assessed. In particular, we modelled the Y-chromosomal genotypes using haploid genotype coding (0,1) to reflect true allele dosage in male individuals, a feature not automatically implemented by all GWAS software.

While the haploid nature of the Y chromosome can increase statistical power for association analyses compared with autosomes due to greater genotype variance, several features complicate statistical inference. In particular, the reduced allelic dosage of the Y chromosome relative to autosomal loci (under the assumption of comparable per-allele effect sizes), combined with the restriction of analyses to male samples, introduces additional challenges in interpreting the results. Moreover, Y-chromosomal association studies test a limited number of common variants, as individual markers often represent extended haplotypes spanning the full length of the male-specific part of the chromosome. Because these variants are highly correlated through shared phylogenetic structure, standard multiple-testing corrections, such as the ones we applied, may be overly conservative. Together, these features imply that the absence of phenome-wide significant associations in our study should not be interpreted as evidence against biologically meaningful effects of Y-chromosomal variation.

Beyond statistical power, the Y chromosome presents distinct challenges for identifying causal variants and biological mechanisms. Extensive linkage across the chromosome renders conventional fine-mapping and colocalization approaches largely inapplicable. To explore potential drivers of association signals, we examined the enrichment of coding variants in each haplogroup, which suggested a possible contribution of *KDM5D* to the I1-related associations observed here. This contrasts with prior work by Eales et al. (2019), which implicated variants in *UTY* in the association between haplogroup I1 and CHD. These differences underscore the difficulty of resolving causal mechanisms on the Y chromosome and highlight the need for complementary functional and evolutionary approaches tailored to its unique genomic architecture.

While the primary focus of this study was on studying SNPs, future studies should benefit from exploring also larger structural and copy number variations^41^. Importantly, the Y chromosome is enriched in repetitive sequences, amplicons, and palindromes^42^, which may contain structural variants affecting gene dosage or regulation. These types of variations in the Y chromosome have been so far mainly linked to infertility but may also impact heritable traits^43^.

In summary, we demonstrate that conducting a PheWAS for Y-chromosomal variation is feasible, and that downstream validation approaches, such as kin-based validation, can help confirm potential genetic associations. Besides providing further support for the link between I1 and CHD, we observed several suggestive signals that warrant follow-up in future studies. Our findings underscore the importance of comprehensively capturing population-specific Y-chromosomal variants to facilitate these efforts. The identified suggestive links to disease risk highlight the need to include the Y chromosome more systematically in larger studies to better understand the male-specific genetic architecture of complex diseases.

## Supporting information

Suppementary tables S1 - S9

## DATA AVAILABILITY

The data used in this study is available through FinnGen upon application (https://www.finngen.fi/en/researchers/accessing). Supplementary material includes Figures S1-S5 and Tables S1-S9. This is an open-access article distributed under the terms of the Creative Commons Attribution 4.0 International License (https://creativecommons.org/licenses/by/4.0/), which permits unrestricted use, distribution, and reproduction in any medium, provided the original work is properly cited.

## ACKNOWLEDGEMENTS

We greatly thank all FinnGen participants as well as the investigators of the FinnGen study. The full FinnGen acknowledgments are provided in the supplemental acknowledgments. We would further like to thank Khujith Rajueni for proofreading of the manuscript. The graphical abstract was created using BioRender (https://www.biorender.com).

## AUTHOR CONTRIBUTION STATEMENT

T.T., J.L. and A.P designed the study. A.P. conducted the analyses. M.P provided statistical assistance. A.L. provided the LOY calls. A.P., J.L., T.T wrote the manuscript. All authors interpreted the results and reviewed the manuscript.

## ETHICAL APPROVAL

Study subjects in FinnGen provided informed consent for biobank research, based on the Finnish Biobank Act. Alternatively, separate research cohorts, collected prior the Finnish Biobank Act came into effect (in September 2013) and start of FinnGen (August 2017), were collected based on study-specific consents and later transferred to the Finnish biobanks after approval by Fimea (Finnish Medicines Agency), the National Supervisory Authority for Welfare and Health. Recruitment protocols followed the biobank protocols approved by Fimea. The Coordinating Ethics Committee of the Hospital District of Helsinki and Uusimaa (HUS) statement number for the FinnGen study is Nr HUS/990/2017. The FinnGen study is approved by Finnish Institute for Health and Welfare (permit numbers: THL/2031/6.02.00/2017, THL/1101/5.05.00/2017, THL/341/6.02.00/2018, THL/2222/6.02.00/2018, THL/283/6.02.00/2019, THL/1721/5.05.00/2019 and THL/1524/5.05.00/2020), Digital and population data service agency (permit numbers: VRK43431/2017-3, VRK/6909/2018-3, VRK/4415/2019-3), the Social Insurance Institution (permit numbers: KELA 58/522/2017, KELA 131/522/2018, KELA 70/522/2019, KELA 98/522/2019, KELA 134/522/2019, KELA 138/522/2019, KELA 2/522/2020, KELA 16/522/2020), Findata permit numbers THL/2364/14.02/2020, THL/4055/14.06.00/2020, THL/3433/14.06.00/2020, THL/4432/14.06/2020, THL/5189/14.06/2020, THL/5894/14.06.00/2020, THL/6619/14.06.00/2020, THL/209/14.06.00/2021, THL/688/14.06.00/2021, THL/1284/14.06.00/2021, THL/1965/14.06.00/2021, THL/5546/14.02.00/2020, THL/2658/14.06.00/2021, THL/4235/14.06.00/2021, Statistics Finland (permit numbers: TK-53-1041-17 and TK/143/07.03.00/2020 (earlier TK-53-90-20) TK/1735/07.03.00/2021, TK/3112/07.03.00/2021) and Finnish Registry for Kidney Diseases permission/extract from the meeting minutes on 4th July 2019. The Biobank Access Decisions for FinnGen samples and data utilized in FinnGen Data Freeze 12 include: THL Biobank BB2017_55, BB2017_111, BB2018_19, BB_2018_34, BB_2018_67, BB2018_71, BB2019_7, BB2019_8, BB2019_26, BB2020_1, BB2021_65, Finnish Red Cross Blood Service Biobank 7.12.2017, Helsinki Biobank HUS/359/2017, HUS/248/2020, HUS/430/2021 §28, §29, HUS/150/2022 §12, §13, §14, §15, §16, §17, §18, §23, §58, §59, HUS/128/2023 §18, Auria Biobank AB17-5154 and amendment #1 (August 17 2020) and amendments BB_2021-0140, BB_2021-0156 (August 26 2021, Feb 2 2022), BB_2021-0169, BB_2021-0179, BB_2021-0161, AB20-5926 and amendment #1 (April 23 2020) and it’s modifications (Sep 22 2021), BB_2022-0262, BB_2022-0256, Biobank Borealis of Northern Finland_2017_1013, 2021_5010, 2021_5010 Amendment, 2021_5018, 2021_5018 Amendment, 2021_5015, 2021_5015 Amendment, 2021_5015 Amendment_2, 2021_5023, 2021_5023 Amendment, 2021_5023 Amendment_2, 2021_5017, 2021_5017 Amendment, 2022_6001, 2022_6001 Amendment, 2022_6006 Amendment, 2022_6006 Amendment, 2022_6006 Amendment_2, BB22-0067, 2022_0262, 2022_0262 Amendment, Biobank of Eastern Finland 1186/2018 and amendment 22§/2020, 53§/2021, 13§/2022, 14§/2022, 15§/2022, 27§/2022, 28§/2022, 29§/2022, 33§/2022, 35§/2022, 36§/2022, 37§/2022, 39§/2022, 7§/2023, 32§/2023, 33§/2023, 34§/2023, 35§/2023, 36§/2023, 37§/2023, 38§/2023, 39§/2023, 40§/2023, 41§/2023, Finnish Clinical Biobank Tampere MH0004 and amendments (21.02.2020 & 06.10.2020), BB2021-0140 8§/2021, 9§/2021, §9/2022, §10/2022, §12/2022, 13§/2022, §20/2022, §21/2022, §22/2022, §23/2022, 28§/2022, 29§/2022, 30§/2022, 31§/2022, 32§/2022, 38§/2022, 40§/2022, 42§/2022, 1§/2023, Central Finland Biobank 1-2017, BB_2021-0161, BB_2021-0169, BB_2021-0179, BB_2021-0170, BB_2022-0256, BB_2022-0262, BB22-0067, Decision allowing to continue data processing until 31st Aug 2024 for projects: BB_2021-0179, BB22-0067,BB_2022-0262, BB_2021-0170, BB_2021-0164, BB_2021-0161, and BB_2021-0169, and Terveystalo Biobank STB 2018001 and amendment 25th Aug 2020, Finnish Hematological Registry and Clinical Biobank decision 18th June 2021, Arctic biobank P0844: ARC_2021_1001.

## REFERENCES

1 Abdellaoui A, Yengo L, Verweij KJH, Visscher PM. 15 years of GWAS discovery: Realizing the promise. The American Journal of Human Genetics 2023; 110: 179–194.

2 Parker K, Erzurumluoglu AM, Rodriguez S. The Y Chromosome: A Complex Locus for Genetic Analyses of Complex Human Traits. [Review]. Genes (Basel) 2020; 11. doi:10.3390/genes11111273.

3 Sun L, Wang Z, Lu T, Manolio TA, Paterson AD. eXclusionarY: 10 years later, where are the sex chromosomes in GWASs? Am J Hum Genet 2023; 110: 903–912.

4 Sollis E et al. The NHGRI-EBI GWAS Catalog: knowledgebase and deposition resource. Nucleic Acids Research 2023; 51: D977–D985.

5 Bruhn-Olszewska B, Markljung E, Rychlicka-Buniowska E, Sarkisyan D, Filipowicz N, Dumanski JP. The effects of loss of Y chromosome on male health. Nat Rev Genet 2025; 26: 320–335.

6 Godfrey AK et al. Quantitative analysis of Y-Chromosome gene expression across 36 human tissues. Genome Res 2020; 30: 860–873.

7 Bellott DW et al. Mammalian Y chromosomes retain widely expressed dosage-sensitive regulators. Nature 2014; 508: 494–499.

8 Lu C et al. Y chromosome haplogroups based genome-wide association study pinpoints revelation for interactions on non-obstructive azoospermia. Sci Rep 2016; 6: 33363.

9 Patel R, Khalifa AO, Isali I, Shukla S. Prostate cancer susceptibility and growth linked to Y chromosome genes. Front Biosci (Elite Ed) 2018; 10: 423–436.

10 Kim W et al. Lack of Association between Y-Chromosomal Haplogroups and Prostate Cancer in the Korean Population. PLoS ONE 2007; 2: e172.

11 Howe LJ, Erzurumluoglu AM, Davey Smith G, Rodriguez S, Stergiakouli E. Y Chromosome, Mitochondrial DNA and Childhood Behavioural Traits. Sci Rep 2017; 7: 11655.

12 Sezgin E et al. Association of Y chromosome haplogroup I with HIV progression, and HAART outcome. Hum Genet 2009; 125: 281.

13 Serajee FJ, Mahbubul Huq AHM. Association of Y chromosome haplotypes with autism. J Child Neurol 2009; 24: 1258–1261.

14 Kim HJ, Jin HJ. Lack of association between the Y chromosome haplogroups and attention deficit hyperactivity disorder (ADHD) in Korean boys. Gene 2023; 850: 146954.

15 Charchar FJ et al. Inheritance of coronary artery disease in men: an analysis of the role of the Y chromosome. The Lancet 2012; 379: 915–922.

16 Eales JM et al. Human Y Chromosome Exerts Pleiotropic Effects on Susceptibility to Atherosclerosis. Arterioscler Thromb Vasc Biol 2019; 39: 2386–2401.

17 Timmers PRHJ, Wilson JF. Limited Effect of Y Chromosome Variation on Coronary Artery Disease and Mortality in UK Biobank-Brief Report. Arterioscler Thromb Vasc Biol 2022; 42: 1198–1206.

18 Sato G et al. Genetic regulation across germline and somatic variation on the Y chromosome contributes to type 2 diabetes. Nat Med 2026. doi:10.1038/s41591-026-04213-z.

19 Kurki MI et al. FinnGen provides genetic insights from a well-phenotyped isolated population. Nature 2023; 613: 508–518.

20 Poznik GD. Identifying Y-chromosome haplogroups in arbitrarily large samples of sequenced or genotyped men. Genetics, 2016 doi:10.1101/088716.

21 Manichaikul A, Mychaleckyj JC, Rich SS, Daly K, Sale M, Chen W-M. Robust relationship inference in genome-wide association studies. Bioinformatics 2010; 26: 2867–2873.

22. https://www.yfull.com/tree/. YFull v10.01. https://www.yfull.com/tree/ (accessed 1 May2024).

23. International Society of Genetic Genealogy (ISOGG) v15.73. https://isogg.org (accessed 11 Jan 2022).

24. FinnGen clinical endpoints DF12. https://www.finngen.fi/en/researchers/clinical-endpoints (accessed 11 Dec 2025).

25 Loh P-R et al. Insights into clonal haematopoiesis from 8,342 mosaic chromosomal alterations. Nature 2018; 559: 350–355.

26 Loh P-R, Genovese G, McCarroll SA. Monogenic and polygenic inheritance become instruments for clonal selection. Nature 2020; 584: 136–141.

27 Koskela JT et al. Genetic variant in SPDL1 reveals novel mechanism linking pulmonary fibrosis risk and cancer protection. 2021. doi:10.1101/2021.05.07.21255988.

28 Zekavat SM et al. Hematopoietic mosaic chromosomal alterations increase the risk for diverse types of infection. Nat Med 2021; 27: 1012–1024.

29. R Core Team. R: A Language and Environment for Statistical Computing. 2025.https://www.R-project.org/.

30 Lenth RV, Piaskowski J. emmeans: Estimated Marginal Means, aka Least-Squares Means. 2017; : 2.0.1.

31 Mbatchou J et al. Computationally efficient whole-genome regression for quantitative and binary traits. Nat Genet 2021; 53: 1097–1103.

32 Wacholder S et al. The Kin-Cohort Study for Estimating Penetrance. American Journal of Epidemiology 1998; 148: 623–630.

33. Pirinen M. linemodels: clustering effects based on linear relationships. Bioinformatics 2023; 39: btad115.

34 Preussner A, Leinonen J, Riikonen J, Pirinen M, Tukiainen T. Y chromosome sequencing data suggest dual paths of haplogroup N1a1 into Finland. Eur J Hum Genet 2025; 33: 89–97.

35. THL:n sairastavuus-indeksi 2019. https://www.terveytemme.fi/sairastavuusindeksi/ (accessed 22 Sept 2025).

36 Kerminen S et al. Geographic Variation and Bias in the Polygenic Scores of Complex Diseases and Traits in Finland. Am J Hum Genet 2019; 104: 1169–1181.

37 Bachtrog D. Y-chromosome evolution: emerging insights into processes of Y-chromosome degeneration. Nat Rev Genet 2013; 14: 113–124.

38 Karczewski KJ et al. The mutational constraint spectrum quantified from variation in 141,456 humans. Nature 2020; 581: 434–443.

39 Sano S et al. Hematopoietic loss of Y chromosome leads to cardiac fibrosis and heart failure mortality. Science 2022; 377: 292–297.

40 Siegel RL, Giaquinto AN, Jemal A. Cancer statistics, 2024. CA A Cancer J Clinicians 2024; **74**: 12–49.

41 Hallast P et al. Assembly of 43 human Y chromosomes reveals extensive complexity and variation. Nature 2023; 621: 355–364.

42 Skaletsky H et al. The male-specific region of the human Y chromosome is a mosaic of discrete sequence classes. Nature 2003; 423: 825–837.

43 Colaco S, Modi D. Consequences of Y chromosome microdeletions beyond male infertility. J Assist Reprod Genet 2019; 36: 1329–1337.

