## Supplementary material and figures S1 - S5 for "Investigating the Y chromosome in complex disease: Phenome-wide scan across 104,334 Finnish men"

### MATERIALS AND METHODS

#### Quality control

For this study, we utilized the FinnGen project[1] release 12 data. Although the majority of the samples in FinnGen have been genotyped using two different arrays custom-designed for the project (ThermoFisher Axiom custom arrays v1 and v2)[2], there are several smaller sub-cohorts included the data set where the Y-chromosomal marker coverage as well as the purpose of the sample collection varies. Besides excluding the so-called legacy samples that consist of cohorts genotyped prior the FinnGen study (N=19,757), we excluded samples genotyped on FinnGen custom array v1 (N=47,199), due to the lack of markers for Finnish-enriched Y-chromosomal haplogroups N1a1, I1 and R1a.

#### Haplogroup-defining SNPs

We used the YFull[3] and ISOGG[4] databases to annotate the haplogroup-defining status of the SNPs in the data. Of note, a notable proportion of the variants discarded from the analyses were non-polymorphic among the samples (N=276; 51%) (especially haplogroup R1b markers), reflecting the western-European bias in the genotyping chip design. Two of the 60 SNPs passing quality control had no known annotations and were not detected in gnomAD[5] or earlier Finnish data[6], despite their high frequency in FinnGen (chrY:11794009 A>T, AF=0.27 and chrY:12487008 C>T, AF=0.89). We therefore considered these as likely false genotypes and removed them from the dataset. One SNP lacked an annotation in YFull or ISOGG (chrY:12225000 A>T, AF=0.26), but had previously been observed in Finnish sequencing data clustering within haplogroup I1[6]. Accordingly, we assigned this variant to haplogroup I1.

#### Exome sequencing data QC and annotation

The reads were aligned to GRCh38, and genotypes were called at the Broad Institute as part of the gnomAD v4[5]. Before quality control, we had 11,209 male samples (non-overlapping with the samples used for the PheWAS) with WES data available for the Y chromosome. We filtered the data by removing genotype calls with DP < 10 or GQ < 20, variants having a mean DP < 10 across all samples and kept only variants annotated with a "PASS" tag in gnomAD[5]. Similarly to the genotyping data, we removed samples with non-Finnish genetic ancestry based on PCA, samples with Klinefelter syndrome, and samples without covariate data, yielding in 9,736 samples. Among this sample set, we kept polymorphic variants, resulting in 300 variants (23 indels, 277 SNPs). The variants were annotated using VEP (version 113)[7] with the additional parameters: CADD score (yes), restrict results (one selected consequence per allele). In addition to the existing VEP annotations, we defined a category of loss of function (LoF), which included frameshift, splice donor, stop gained, splice acceptor variants.

### Phenotype selection

We further included 13 quantitative endpoints including height, weight, number of offspring, and common laboratory measurements into the analyses. These included measurements of hemoglobin, hemoglobin A1C, glucose, HDL, LDL, triglycerides, alanine aminotransferase, aspartate transaminase, bilirubin, creatine. From laboratory measurements we removed outliers falling into these ranges: LDL < 0 or > 20 (N=14); HDL > 15 (N=15); glucose < 1 or > 100 (N=4); creatine > 10,000 (N=1). All the quantitative phenotypes except for number of offspring were normalized using inverse normal transformation.

### Statistical power calculations

In addition, statistical power analyses were performed for each haplogroup separately by computing statistical power for a range of 500 theoretical effect sizes for a given haplogroup and trait. To estimate the smallest effect size for which we had adequate power, we calculated the beta which was required to achieve ≥80% power for each haplogroup under different sample size scenarios. To estimate power for binary traits, this was done using three different number of cases (100, 1,000, and 10,000) with a total sample size of N = 100,000 and for quantitative traits, calculations were performed with N = 100,000. Using these parameters, power was calculated for a set of betas in R (version 4.5.0)[8] for binary traits as

$$
\begin{aligned}
&power.bin = function(p, \beta, f, n.cases, n.controls)\{\\
&\quad q = qchisq(p, df = 1, ncp = 0, lower = F)\\
&\quad ncps = \frac{n.cases * n.controls}{n.cases + n.controls} * f * (1-f) * \beta^2\\
&\quad power = pchisq(q, df = 1, ncp = ncps, lower = F)\\
&\quad return(power)\\
&\}
\end{aligned}
$$

and for quantitative traits as

$$
\begin{aligned}
&power.quant = function(p, \beta, f, n)\{\\
&\quad h^2 = f * (1-f) * \beta^2\\
&\quad q = qchisq(p, df = 1, ncp = 0, lower = F)\\
&\quad power = pchisq(q, df = 1, ncp = n * h^2, lower = F)
\end{aligned}
$$

$$return(power)$$

$$\}$$

where p is the PheWAS p-value threshold, β the given effect size(s) under allele coding {0,1}, f the minor allele frequency, n.cases the number of cases, n.controls the number of controls and n the total number of samples.

### Kin-based validation - Effect size comparison between first degree relatives

For the kin-based validation analysis, we identified relatives for the N=104,334 samples used for the PheWAS. Using the relatedness analysis done by FinnGen using KING[9] for 520,210 samples, we first identified all samples showing 1st degree relatedness (N=30,245 males, N=23,217 females) with our samples. We then filtered these male and female kin-cohorts by removing closely related samples within the kin-cohorts (≤2nd degree relatives) to avoid any bias in the association analyses, resulting in cohorts of N=24,712 males (N=7,922 brothers, N=7999 fathers, N=8791 sons) and N=19,275 females (N=4,544 daughters, N=2,979 mothers, N=11,761 sisters) for the kin-based validation.

After the association analyses using the kin cohorts, we classified the associations as concordant or discordant by evaluating the posterior probabilities (PP) of the female and male validations under the models M1 and M0 from linemodels[10]. Concordant effects were classified as (Male M1 PP > 0.5 & Female M1 PP < 0.5). Among the discordant effects, we classified the effects as: 1) concordant in females (Female M1 PP > 0.5; Male M1 PP < 0.5), 2) both relatives (Female M1 PP > 0.5; Male M1 PP > 0.5), or 3) neither of the relatives (Female M1 PP < 0.5; Male M1 PP < 0.5). The posterior probability threshold of 0.5 indicates which of the two models is more probable given the data. While this approach is not formally assessing statistical significance, it provides a useful summary of the data given the relatively small sample size and generally low observed PPs (**Table S5**).

### Assessing effects between haplogroup pairs

Since the effect size of a given SNP or haplogroup depends on the reference group used for comparison, that vary by populations, we carried out additional analyses assessing each haplogroup's effect in relation to another. These pairwise analyses are needed to accurately compare the effect of a given haplogroup between populations with divergent haplogroup compositions, such as Finland and the UK (where the Finnish-enriched haplogroup N1a1 is nearly absent). To this end, we selected haplogroups N1a1, I1, R1a and R1b for pairwise comparisons and performed analyses in REGENIE[11] on subsets of samples representing each haplogroup pair (e.g., N1a1 against I1 carriers within one association test), repeating this across all suggestive associations.

### Adjusting for possible CHD confounders

Further to assess whether the association for CHD could be driven by autosomal genetic variation rather than Y-chromosomal variation, we incorporated a polygenic risk score (PRS) for CHD into the analysis (computed by the FinnGen analysis team with PRS-CS[12], using GWAS data[13], standardized to mean=0 and SD=1) into the analysis.

To account for potential geographical confounding, we conducted stratified analyses across distinct regions. First, we assessed whether the effect was consistent across the samples' birth regions (N=20) in Finland by performing separate association analysis within each region and compared the overall effects using a Q-test for heterogeneity. To enhance the statistical power and robustness of the regional effect estimates, we conducted geographically stratified association analyses between northeastern (NE) and southwestern (SW) regions in Finland. The NE and SW regions were defined according to the previously reported autosomal population structure[14]. Lapland, Northern Ostrobothnia, Kainuu, Northern Savonia, Southern Savonia, Northern Karelia, Southern Karelia, ceded Karelia and Central Finland were categorized as NE (N=40,328), whereas Central Ostrobothnia, Ostrobothnia, Southern Ostrobothnia, Satakunta, Pirkanmaa, Southwest Finland, Kanta-Häme, Päijät-Häme, Uusimaa, Kymenlaakso and Åland were categorized as SW (N=62,834).

### REFERENCES


1 Kurki MI *et al.* FinnGen provides genetic insights from a well-phenotyped isolated population. *Nature* 2023; **613**: 508–518.

2 FinnGen genotype data. https://docs.finngen.fi/finngen-data-specifics/red-library-data-individual-level-data/genotype-data/affymetrix-chip-and-its-design (accessed 11 Dec 2025).

3 YFull v10.01. https://www.yfull.com/tree/ (accessed 1 May 2024).

4 International Society of Genetic Genealogy (ISOGG) v15.73. https://isogg.org (accessed 11 Jan 2022).

5 Koenig Z *et al.* A harmonized public resource of deeply sequenced diverse human genomes. *bioRxiv* 2023; : 2023.01.23.525248.

6 Preussner A, Leinonen J, Riikonen J, Pirinen M, Tukiainen T. Y chromosome sequencing data suggest dual paths of haplogroup N1a1 into Finland. *Eur J Hum Genet* 2025; **33**: 89–97.

7 McLaren W *et al.* The Ensembl Variant Effect Predictor. *Genome Biol* 2016; **17**: 122.

8 R Core Team. R: A Language and Environment for Statistical Computing. 2025.https://www.R-project.org/.

9 Manichaikul A, Mychaleckyj JC, Rich SS, Daly K, Sale M, Chen W-M. Robust relationship inference in genome-wide association studies. *Bioinformatics* 2010; **26**: 2867–2873.

10 Pirinen M. linemodels: clustering effects based on linear relationships. *Bioinformatics* 2023; **39**: btad115.

11 Mbatchou J *et al.* Computationally efficient whole-genome regression for quantitative and binary traits. *Nat Genet* 2021; **53**: 1097–1103.

12 Ge T, Chen C-Y, Ni Y, Feng Y-CA, Smoller JW. Polygenic prediction via Bayesian regression and continuous shrinkage priors. *Nat Commun* 2019; **10**: 1776.

13 the CARDIoGRAMplusC4D Consortium. A comprehensive 1000 Genomes–based genome-wide association meta-analysis of coronary artery disease. *Nat Genet* 2015; **47**: 1121–1130.

14 Kerminen S *et al.* Fine-Scale Genetic Structure in Finland. *G3 (Bethesda)* 2017; **7**: 3459–3468.

### FINNGEN SUPPLEMENTARY ACKNOWLEDGEMENTS

We want to acknowledge the participants and investigators of the FinnGen study. The FinnGen project is funded by two grants from Business Finland (HUS 4685/31/2016 and UH 4386/31/2016) and the following industry partners: AbbVie Inc., AstraZeneca UK Ltd, Biogen MA Inc., Bristol Myers Squibb Inc. (and Celgene Corporation & Celgene International II Sàrl), Genentech Inc., Merck Sharp & Dohme LCC, Pfizer Inc., GlaxoSmithKline Intellectual Property Development Ltd., Sanofi US Services Inc., Maze Therapeutics Inc., Johnson&Johnson Innovative Medicine Inc., Novartis AG, Boehringer Ingelheim International GmbH and Bayer AG. Following biobanks are acknowledged for delivering biobank samples to FinnGen: Auria Biobank (www.auria.fi/biopankki), THL Biobank (www.thl.fi/biobank), Helsinki Biobank (www.helsinginbiopankki.fi), Biobank Borealis of Northern Finland (https://www.ppshp.fi/Tutkimus-ja-opetus/Biopankki/Pages/Biobank-Borealis-briefly-in-English.aspx), Finnish Clinical Biobank Tampere (www.tays.fi/en-US/Research_and_development/Finnish_Clinical_Biobank_Tampere), Biobank of Eastern Finland (www.ita-suomenbiopankki.fi/en), Central Finland Biobank (www.ksshp.fi/fi-FI/Potilaalle/Biopankki), Finnish Red Cross Blood Service Biobank (www.veripalvelu.fi/verenluovutus/biopankkitoiminta), Terveystalo Biobank (www.terveystalo.com/fi/Yritystietoa/Terveystalo-Biopankki/Biopankki/) and Arctic Biobank (https://www.oulu.fi/en/university/faculties-and-units/faculty-medicine/northern-finland-birth-cohorts-and-arctic-biobank). All Finnish Biobanks are members of BBMRI.fi infrastructure (https://www.bbmri-eric.eu/national-nodes/finland/). Finnish Biobank Cooperative - FINBB (https://finbb.fi/) is the coordinator of BBMRI-ERIC operations in Finland. The Finnish biobank data can be accessed through the Fingenious® services (https://site.fingenious.fi/en/) managed by FINBB.

### SUPPLEMENTARY FIGURES AND TABLES

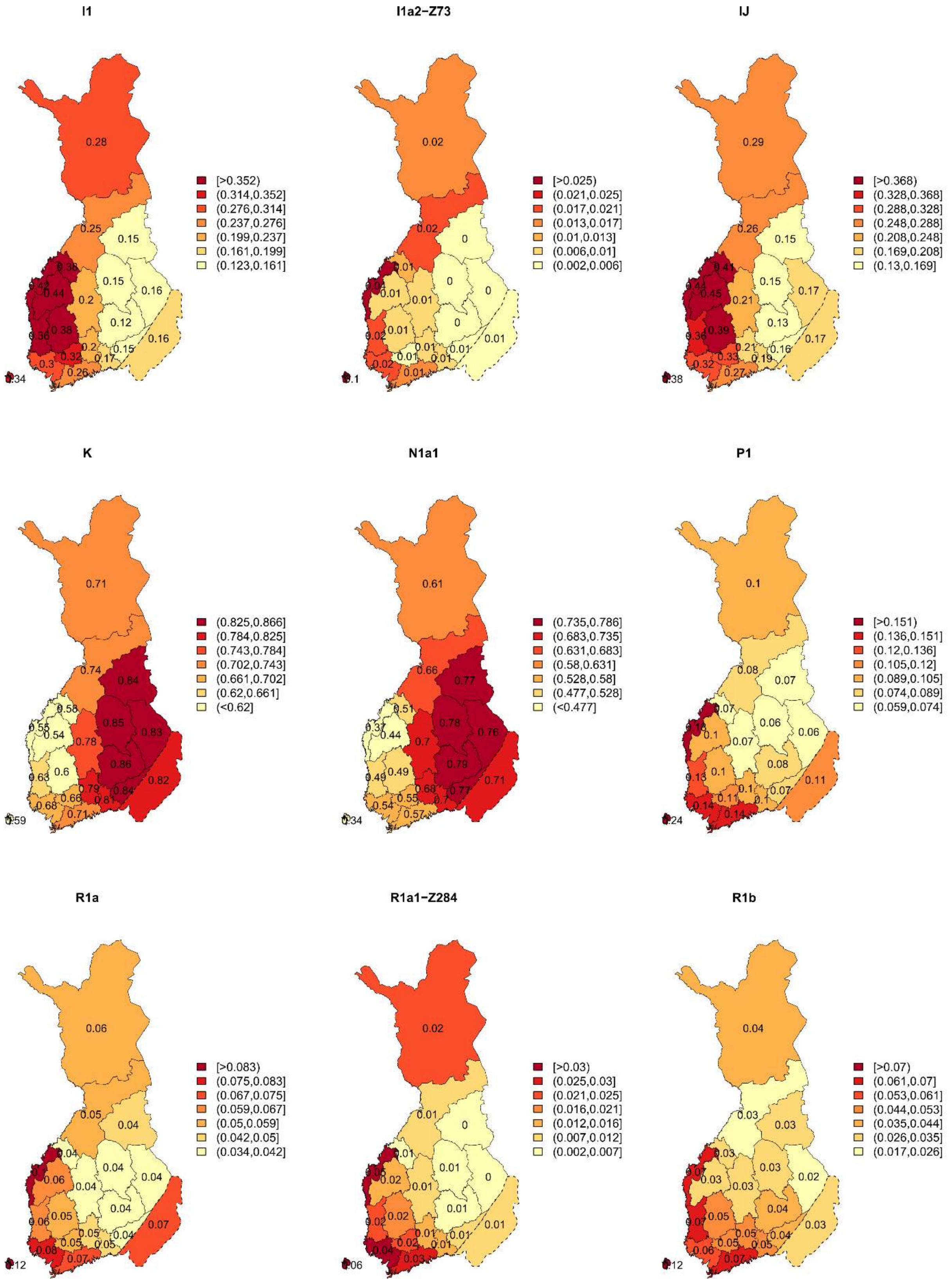


**Figure S1)** Geographical distribution of the nine haplogroups analysed in the PheWAS, shown as regional frequencies among 20 Finnish regions.

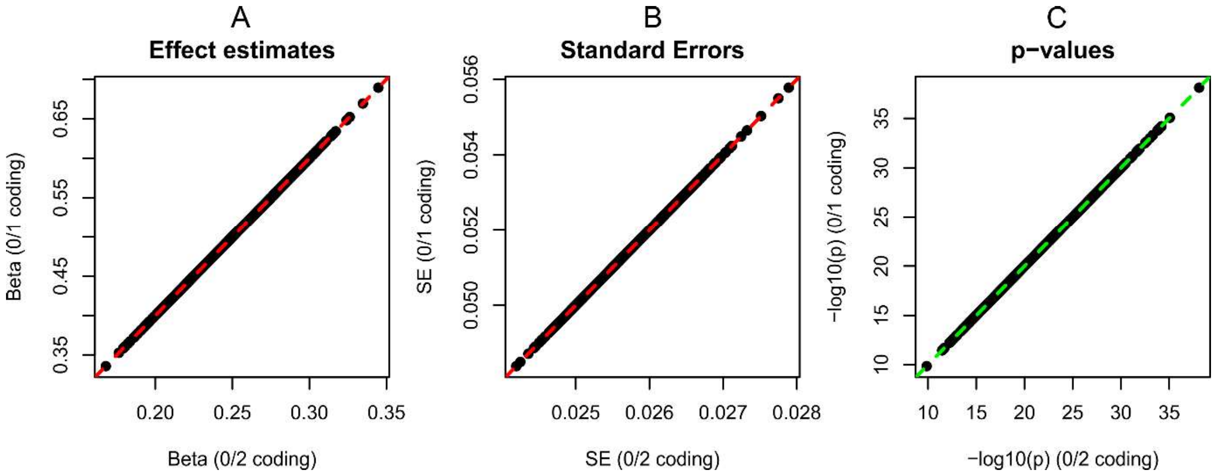


**Figure S2)** Comparison of the haploid genotype coding (0/1) and diploid coding (0/2) for A) effect estimates B) standard errors, and C) p-values. The displayed data were simulated for 100,000 individuals, with genotypes generated under a binomial distribution assuming a minor allele frequency of 0.1. Results are based on 1,000 simulation replicates of linear regression with a continuous phenotype generated according to a linear model with $\beta = 0.5$ and residual variance $\sigma^2 = 1$. The red dashed line corresponds to the reference line $y = 2x$, while the green line corresponds to $y = x$.

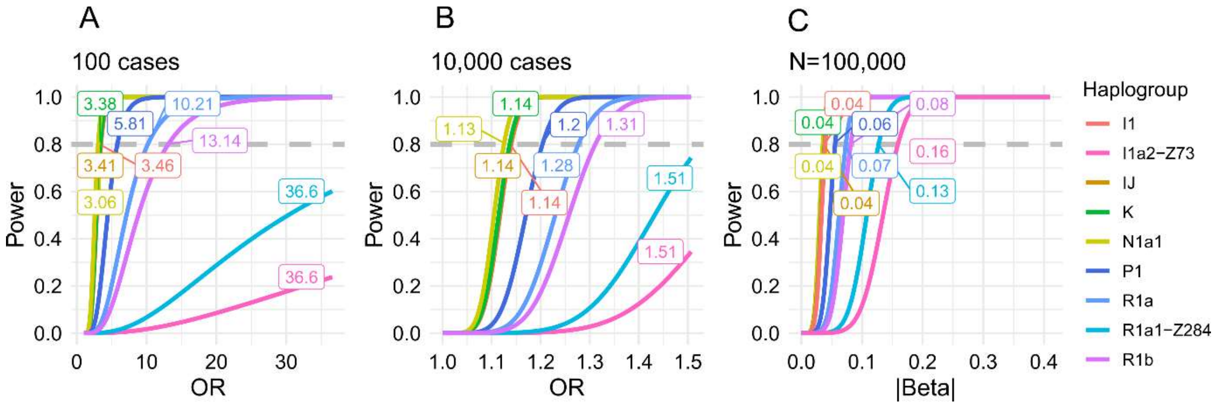


**Figure S3)** Smallest effect sizes that could be reliably detected (80% power) within each haplogroup for traits with A) 100 cases, B) 10,000 cases as well as C) for quantitative traits at p=0.05/9/1,427. Each haplogroup analysed in the PheWAS is annotated with a color as shown in the legend.

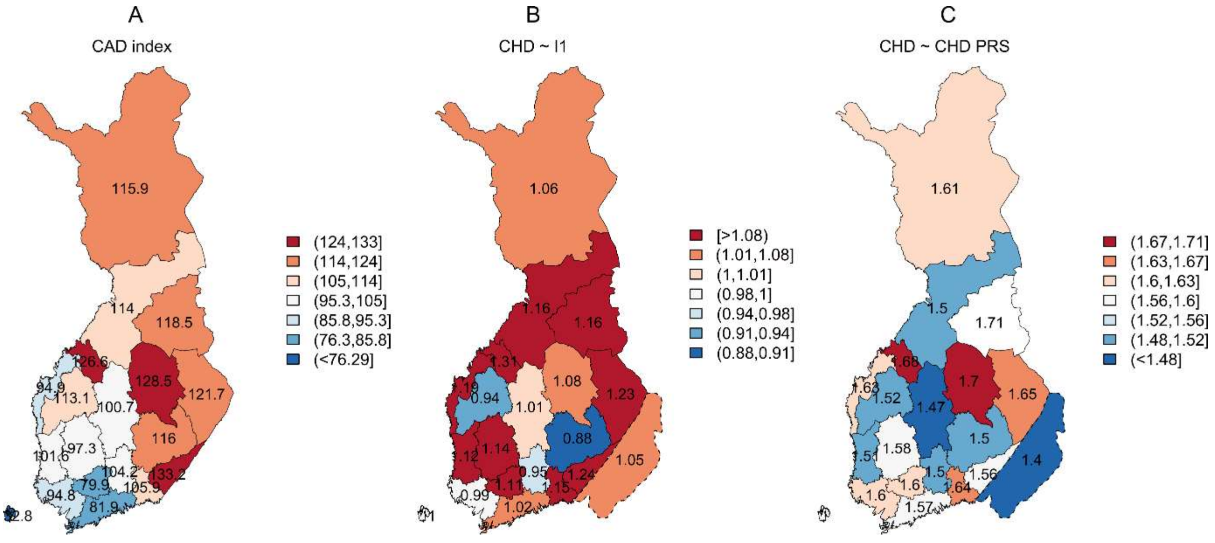


**Figure S4)** Geographical distribution of A) age-adjusted CAD index in Finland from 2017-2019, where the index 100 represents the mean across the country [29]. B) Effect sizes of the association between I1 and CHD, and C) the association between CHD PRS and CHD regionally.

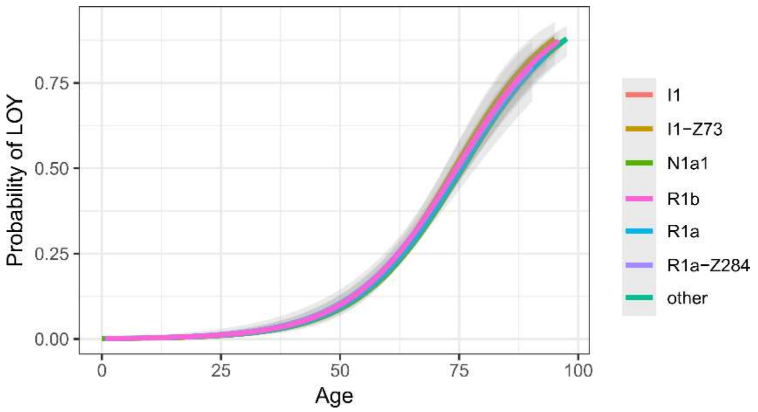


**Figure S5)** Frequency of LOY across age groups by haplogroup. The “other” category includes remaining samples not assigned to the specified haplogroups.
